# Financial Conflicts of Interest among U.S. Physician Authors of 2020 Clinical Practice Guidelines: A Cross-Sectional Study

**DOI:** 10.1101/2022.10.14.22281111

**Authors:** Maryam Mooghali, Laura Glick, Reshma Ramachandran, Joseph S. Ross

**Affiliations:** Section of General Internal Medicine, Department of Internal Medicine, Yale School of Medicine, New Haven, CT, USA; Yale Collaboration for Research Integrity and Transparency (CRIT), Yale School of Medicine, New Haven, CT, USA; Department of Internal Medicine, Yale School of Medicine, Yale University, New Haven, CT, USA; Department of Health Policy and Management, Yale School of Public Health; and Center for Outcomes Research and Evaluation, Yale-New Haven Health System, New Haven, CT, USA

**Keywords:** conflict of interest, industry payments, clinical practice guidelines

## Abstract

**Objective:** To evaluate the prevalence and accuracy of industry-related financial conflict of interest (COI) disclosures among U.S. physician guideline authors

**Design:** Cross-sectional study

**Setting:** Clinical practice guidelines published by the Council of Medical Specialty Societies in 2020

**Participants:** U.S. physician guideline authors

**Main outcome measures:** Financial COI disclosures, both self-reported and determined using Open Payments data

**Results:** Among 270 U.S. physician authors of 20 clinical practice guidelines, 101 (37.4%) disclosed industry-related financial COIs, whereas 199 (73.7%) were found to have received payments from industry when accounting for payments disclosed through Open Payments. The median payments received by authors during the 3-year period was $27,451 (interquartile range [IQR], $1,385-$254,677). Comparing authors’ self-disclosures with Open Payments, 72 (26.7%) of the authors accurately disclosed their financial COIs, including 68 (25.2%) accurately disclosing no financial COIs and 4 (1.5%) accurately disclosing a financial COI. In contrast, 101 (37.4%) disclosed no financial COIs and were found to have received payments from industry, 23 (8.5%) disclosed a financial COI but had underreported payments received from industry, 14 (5.2%) disclosed a financial COI but had overreported payments received from industry, and 60 (22.2%) disclosed a financial COI but were found to have both underreported and overreported payments received from industry. We found that inaccurate COI disclosure was more frequent among professors compared to non-professors (81.9% vs. 63.5%; p<0.001) and among males compared to females (77.7% vs 64.8%; p=0.02). The accuracy of disclosures also varied among medical professional societies (p<0.001).

**Conclusions:** Financial relationships with industry are common among U.S. physician authors of clinical practice guidelines and are often not accurately disclosed. To ensure high-quality guidelines and unbiased recommendations, more effort is needed to minimize existing COIs and improve disclosure accuracy among panel members.

**What is already known on this topic:**

- Clinical practice guidelines are commonly used by physicians and influence patient care decisions
- Financial conflicts of interest among authors of clinical practice guidelines could compromise their integrity

**What this study adds:**

- Financial conflicts of interest are common among U.S. physician authors of clinical practice guidelines and often are not disclosed or disclosed inaccurately
- Although a significant proportion of the monetary value of industry payments received from guideline authors was associated with research activities through institutions, authors were more likely to have undisclosed or underreported COIs for direct payments

**Strength and limitations of this study:**

- Our study included a wide range of contemporary clinical practice guidelines from different professional societies, enhancing relevance and generalizability.
- We were limited to characterizing disclosures only for U.S. physicians.
- We only considered financial COIs with the pharmaceutical and medical device industry.

## INTRODUCTION

Clinical practice guidelines are commonly used by clinicians to inform patient care decisions. The National Academy of Medicine (formerly called the Institute of Medicine) has defined conflict of interest (COI) as “circumstances that create a risk that professional judgments or actions regarding a primary interest will be unduly influenced by a secondary interest” and have the potential to undermine guidelines’ quality, reliability, and integrity, resulting in harm to patients, healthcare professionals, and the healthcare systems.^1,2^ Prior studies have demonstrated an association between guideline authors’ financial COIs with industry and favorable recommendations for their products.^3,4^ Therefore, full disclosure of financial COIs has been mandated by several medical professional societies issuing guidelines, the National Academy of Medicine, and the World Health Organization, emphasizing the importance of making transparent potential COIs among panel members who participate in the development of the clinical practice guidelines.^2,5,6^

Despite increased requirements for guideline authors to have limited COIs and to fully disclose COIs when present, studies have shown high rates of financial relationships among guideline panel members, many of which are undisclosed or underreported.^7-11^ A recent systematic review of nearly 15,000 guideline authors found that 45% reported a financial COI,^7^ however, 32% of authors had undisclosed financial relationships with the industry.^7^ In 2014, data representing payments from industry to U.S.-based physicians was first made available through the Centers for Medicare and Medicaid Services (CMS) Open Payments program, enabling numerous studies comparing disclosures by clinical practice guideline authors to those reported to CMS by manufacturers. However, many of these were conducted for guidelines issued by a single professional society or very soon after the Open Payments program went into effect,^7,12-16^ before physicians may have realized that there would be opportunities for external scrutiny of their disclosures.^17^

Accordingly, our objective was to examine the accuracy of disclosed financial COIs among a more contemporary sample of U.S. physician authors of clinical practice guidelines in 2020. We hypothesized that with the availability of the Open Payments database, most guideline authors would disclose their COIs accurately and expected modest differences in the disclosure of financial COIs among medical professional societies. We also evaluated the scope and nature of the payments received by U.S. physician guideline authors.

## METHODS

This cross-sectional study examined the prevalence and monetary value of financial COIs for authors of guidelines published in 2020 that were issued by any eligible member organization of the Council of Medical Specialty Societies (CMSS). The study also examined the concordance of COIs self-reported by the guideline authors and those listed for each author with a profile on the CMS Open Payments program database. Financial COIs were determined using the publicly available guideline materials and the Open Payments program database.^18^ Since publicly available nonclinical datasets were used, informed consent and institutional review board approval were not required. Patients or the public were not involved in the design, or conduct, or reporting, or dissemination plans of our research. Findings were reported according to the STROBE (Strengthening the Reporting of Observational studies in Epidemiology) guidelines.^19^

### Sample

We identified one guideline from each of the medical professional societies that were member organizations of the CMSS in 2020.^20^ For societies with multiple clinical practice guidelines, we chose the one with the largest number of authors. We included guidelines that were authored by multiple societies if all were members of the CMSS. We excluded systematic review documents that were not endorsed by the associated society as official guidelines. For all authors, we recorded the name, gender, degree, academic rank, country of practice, and whether they were panel chairs of eligible guidelines. We determined the rank (as of 2020) and gender of each author using their academic profile webpages. If the gender or associated pronoun was not available on the institution profile page, we used Google searches to identify gender and matched them with available profile photos. Authors from outside the United States and those who were not physicians (e.g., PhDs) were excluded from the analysis, as Open Payments, as of 2020 under the Physician Payments Sunshine Act, only required disclosure of payments from industry to U.S. physicians and academic medical centers.^21^

### Main Outcome measure

We searched the main documents and supplementary files for each guideline and collected the industry-related declared financial COIs (collected by MM and LG). Financial disclosures related to payments from foundations, medical professional societies, academic institutions, and governmental entities were excluded. Industry payments over the prior three years were determined from the Open Payments database (in alignment with the International Committee of Medical Journal Editors’ (ICMJE) recommended timespan for disclosing any potential COIs).^22^ To facilitate data collection, we collected information on all payments from January 1, 2017 to December 31, 2019 for all guidelines accepted for publication before January 2020 or published before March 2020. For the remaining guidelines, we collected information on all payments over the three-year period before acceptance for publication. If the acceptance date was not available, we assumed that the guideline was accepted three months before the publication date.

Financial COIs were defined as any payments received by a guideline author from pharmaceutical or medical device companies. The payments included research funding and general payments, as categorized by CMS.^23^ Research funding could be paid either directly to the recipient (“Research Payment”) or through a research institution or entity where the recipient was a principal investigator (“Associated Research Funding”). General payments covered fees for non-research activities such as consulting, honoraria, royalty or license, education, gifts, travel and lodging, and food and beverage. Ownership and investment interest of authors were excluded. ^24^ We categorized payments as either “Direct Payment”, including general payments and direct research payments, and “Associated Research Funding”, which were received through a research organization. Data collection from Open Payments was done in May and June 2022.

For each guideline author, we first confirmed their identity by matching their name, specialty, and practice location reported on their Open Payment profile with their information in the guidelines. Next, we compared the data collected from Open Payments with authors’ self-disclosed COIs. If the source of payment found on Open Payments matched with the declared COI, that payment was considered as a disclosed COI. Otherwise, it was recorded as an undisclosed COI. Total COIs were calculated by adding the disclosed and undisclosed COIs.

We categorized the status of financial COIs into the following groups: (1) undeclared in the guideline and no payments found on Open Payments (accurate disclosure of no financial COIs), (2) undeclared in the guideline but payments found on Open Payments, (3) disclosure of payments in the guideline and no additional payments found on Open Payments (accurate disclosure of financial COIs), (4) disclosure of payments in the guideline but additional payments found on Open Payments (underreporting), (5) disclosure of payments in the guideline but not all payments were found on Open Payments (overreporting), (6) disclosure of payments in the guidelines, but both additional payments were found and not all disclosed payments were found on Open Payments (underreporting and overreporting).

### Patient and Public Involvement

None

### Statistical Analysis

We reported the prevalence and accuracy of disclosure of financial COIs, as well as the types and amounts of compensation received by all guideline authors. We also examined whether there were any associations between the accuracy of COI disclosure with gender, rank, role as panel chair, and medical professional society. We analyzed the differences between each group by using a two-sided, chi-squared test. A p-value<0.05 was considered statistically significant. Data were recorded and categorized in Microsoft Excel software, 2018 (Microsoft Corp). We used JMP Pro, Version 16.2 (SAS Institute Inc) for conducting the chi-squared tests.

## RESULTS

### Sample characteristics

A total of 20 guidelines were included in our study, listed in Supplemental Table 1. All guidelines were issued by a medical professional society with a COI policy for panel members, and all the guidelines provided an opportunity for authors to publicly disclose their financial COIs. The median number of guideline authors was 16 (interquartile range [IQR], 9-24). A total of 371 individuals were listed as authors of the 20 guidelines, of which 101 (27.2%) were based outside the U.S and/or did not have an MD/DO/MBBS degree. Thus, 270 authors, representing 267 unique individuals, were included in the analysis; 3 individuals were listed as authors of two guidelines. Of the 270 authors included in the analysis, 177 (65.6%) were male, 144 (53.3%) were of the professor rank, and 22 (8.1%) were panel chairs. Additional characteristics of total 371 authors and the 270 included authors are summarized in Supplemental Table 2 and Table 1, respectively.

**Table 1.**
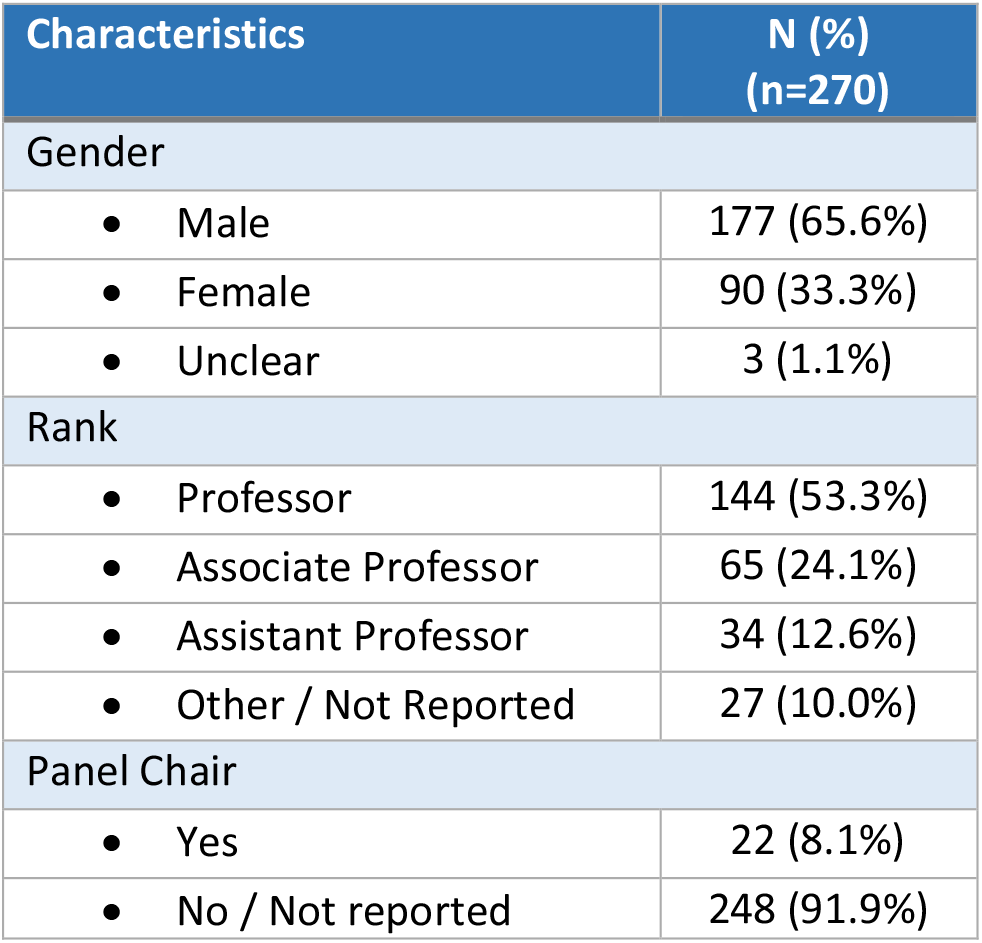
Characteristics of U.S. Physician Authors of 2020 Clinical Practice Guidelines published by the Council of Medical Specialty Societies.

### Prevalence of financial COIs

Of the 270 panel members, 101 (37.4%) declared financial COIs and 169 (62.6%) did not declare any financial COIs. However, when accounting for disclosures listed on Open Payments, 199 (73.7%) were found to have received payments from industry. Authors with COI comprised the minority of the panel for only 5 (25.0%) guidelines. Among the 22 panel chairs, 7 (31.8%) declared financial COIs. However, when accounting for disclosures listed on Open Payments, 18 (81.8%) had financial COIs, none of which disclosed their COI accurately.

Comparing authors’ self-disclosures with Open Payments, 72 (26.7%) of the authors accurately disclosed their financial COIs, including 68 (25.2%) accurately disclosing no financial COIs and 4 (1.5%) accurately disclosing a financial COI. In contrast, 101 (37.4%) disclosed no financial COIs and were found to have received payments from industry, 23 (8.5%) disclosed a financial COI but had underreported all payments received from industry, 14 (5.2%) disclosed a financial COI but had overreported payments received from industry, and 60 (22.2%) disclosed a financial COI but were found to have both underreported and overreported payments received from industry (Table 2).

**Table 2.** Financial Conflict of Interest Disclosures among U.S. Physician Authors of 2020 Clinical Practice Guidelines.

|  | N (%)<br>(n=270) |
| --- | --- |
| Undeclared in the guideline and no payments found on Open Payments (accurate disclosure of no financial COIs) | 68 (25.2%) |
| Undeclared in the guideline but payments found on Open Payments | 101 (37.4%) |
| Disclosure of payments in the guideline and no additional payments found on Open Payments (accurate disclosure of financial COIs) | 4 (1.5%) |
| Disclosure of payments in the guideline but additional payments found on Open Payments (underreporting) | 23 (8.5%) |
| Disclosure of payments in the guideline but not all payments were found on Open Payments (overreporting) | 14 (5.2%) |
| Disclosure of payments in the guidelines, but both additional payments were found and not all disclosed payments were found on Open Payments (underreporting and overreporting) | 60 (22.2%) |

Abbreviations: COI = conflict of interest.

### Conflict of interest by authors’ characteristics and societies

Inaccurate disclosures of financial COIs were more common by professors compared with non-professors or those with unavailable rank (81.9% vs. 63.5%; p<0.001) and by male authors compared with female authors (77.7% vs. 64.8%; p=0.02). Furthermore, the accuracy of COIs reported among the medical professional societies statistically differed (p<0.001), as the American Society of Colon and Rectal Surgeons (ACSRS) and Society for Vascular Surgery (SVS) had the highest inaccuracy rates (100%), whereas the American College of Physicians (ACP) had the lowest inaccuracy rate (25.0%). We found no statistically significant difference in the accuracy of COIs reported among panel chairs compared with other panel members (Table 3).

**Table 3.**
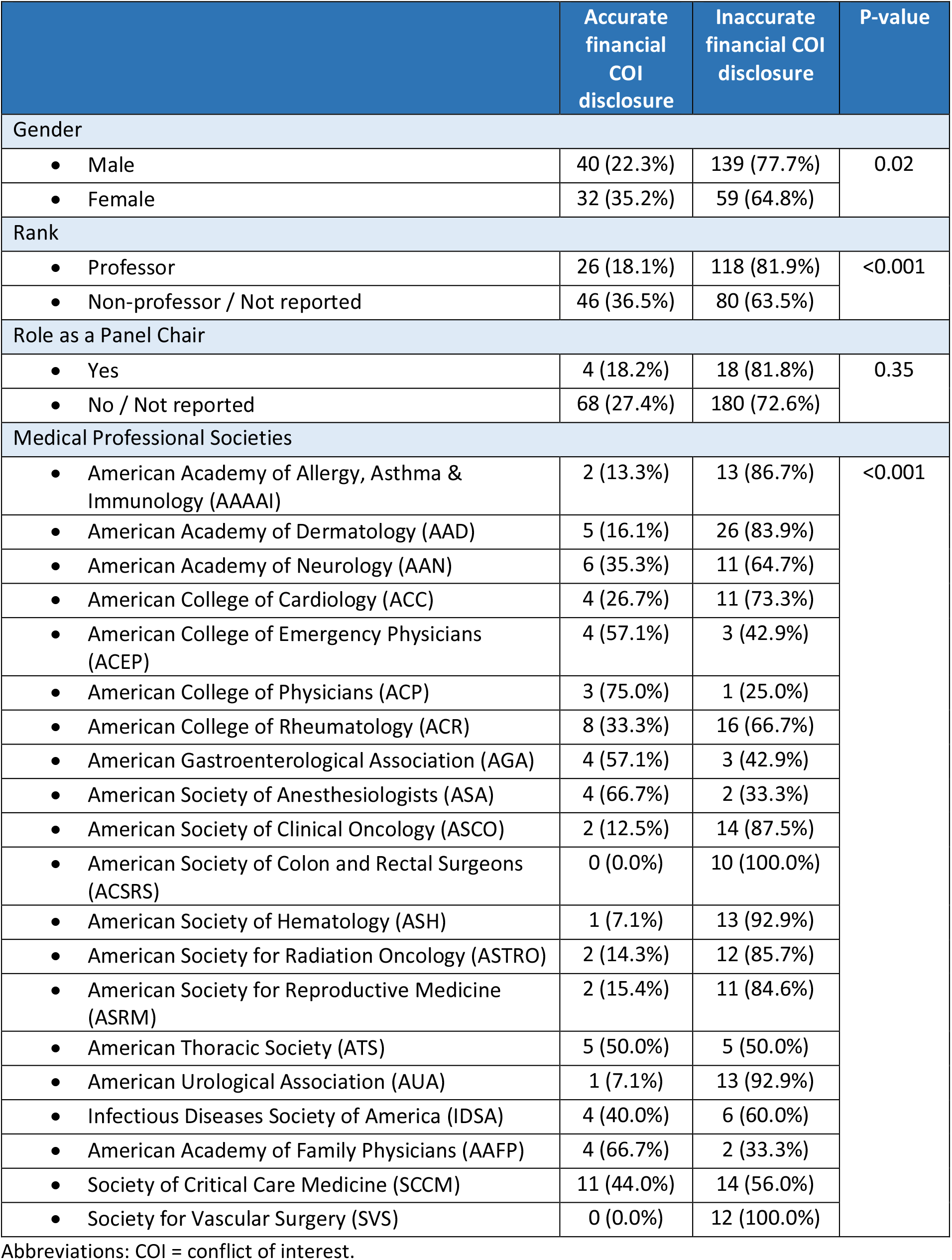
Accuracy of Financial Conflict of Interest Disclosures among U.S. Physician Authors of 2020 Clinical Practice Guidelines, Stratified by Author and Guideline Characteristics.

#### Authors with identified COIs on Open Payments

Based on the search conducted on Open Payments, 199 authors had financial COIs listed on the database, with the median 3-year payments of $27,451 (IQR, $1,385-$254,677). The values of total and undisclosed COIs were $98,716,681 and $23,976,655, respectively. Over 80% of COIs were received as Associated Research Funding (median $154 [IQR, $0-$212,932]), and the median value of general payments and research payments received directly by physicians were $5,487 (IQR, $344-$48,834) and $0 ($0-$770), respectively (Table 4).

**Table 4.**
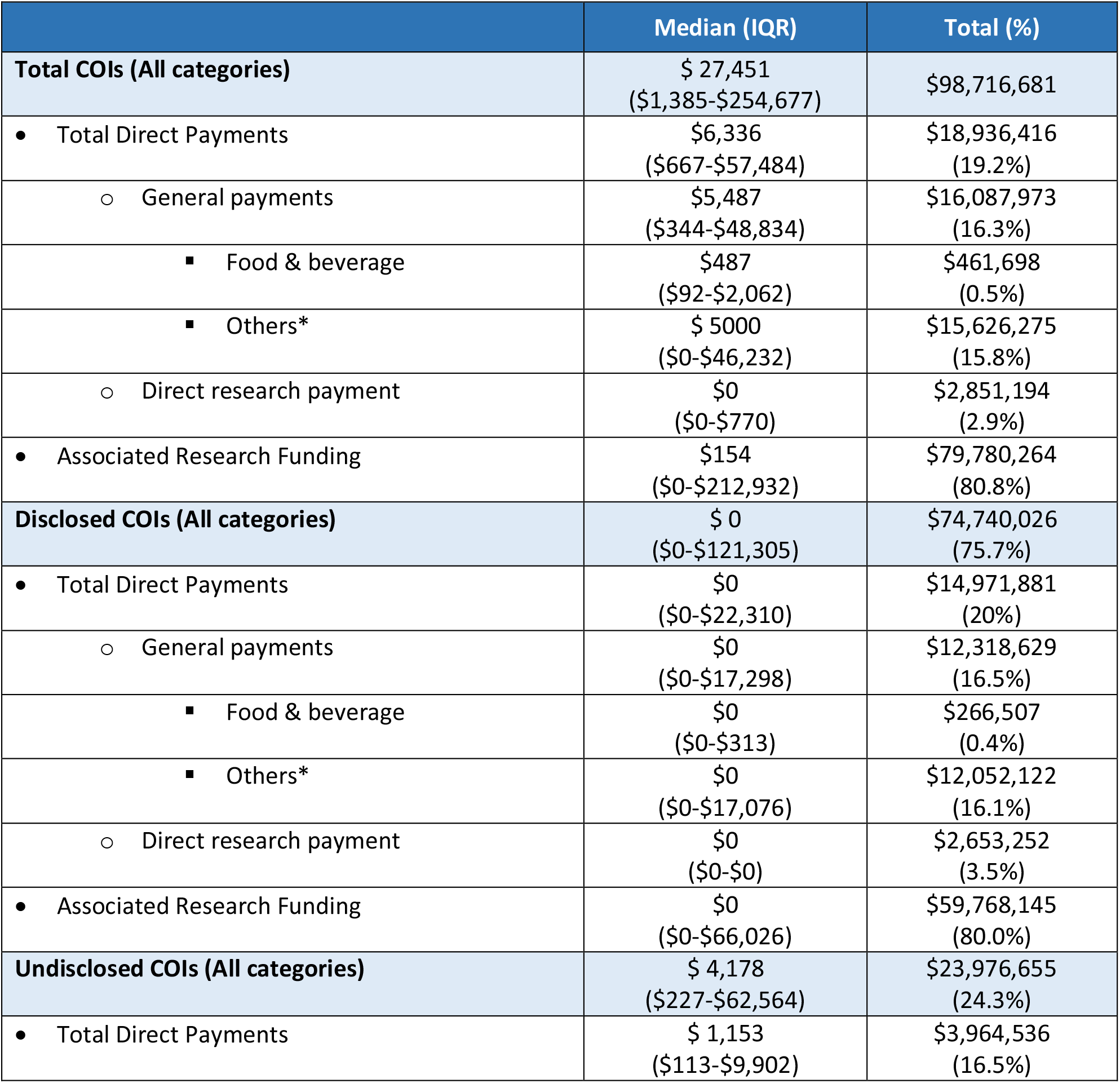
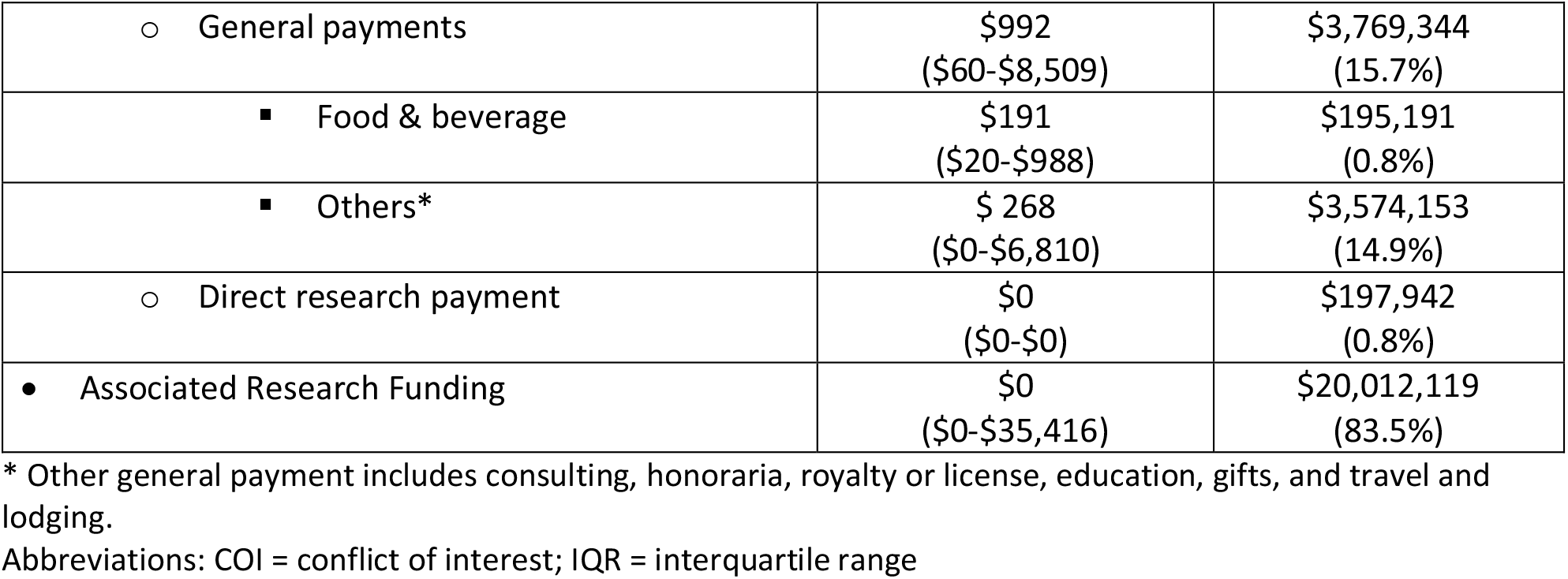
Monetary Value of Financial Conflict of Interests among U.S. Physician Authors of 2020 Clinical Practice Guidelines.

|  | Median (IQR) | Total (%) |
| --- | --- | --- |
| **Total COIs (All categories)** | \$ 27,451<br>(\$1,385-\$254,677) | \$98,716,681 |
| • Total Direct Payments | \$6,336<br>(\$667-\$57,484) | \$18,936,416<br>(19.2%) |
| ○ General payments | \$5,487<br>(\$344-\$48,834) | \$16,087,973<br>(16.3%) |
| ▪ Food & beverage | \$487<br>(\$92-\$2,062) | \$461,698<br>(0.5%) |
| ▪ Others* | \$ 5000<br>(\$0-\$46,232) | \$15,626,275<br>(15.8%) |
| ○ Direct research payment | \$0<br>(\$0-\$770) | \$2,851,194<br>(2.9%) |
| • Associated Research Funding | \$154<br>(\$0-\$212,932) | \$79,780,264<br>(80.8%) |
| **Disclosed COIs (All categories)** | \$ 0<br>(\$0-\$121,305) | \$74,740,026<br>(75.7%) |
| • Total Direct Payments | \$0<br>(\$0-\$22,310) | \$14,971,881<br>(20%) |
| ○ General payments | \$0<br>(\$0-\$17,298) | \$12,318,629<br>(16.5%) |
| ▪ Food & beverage | \$0<br>(\$0-\$313) | \$266,507<br>(0.4%) |
| ▪ Others* | \$0<br>(\$0-\$17,076) | \$12,052,122<br>(16.1%) |
| ○ Direct research payment | \$0<br>(\$0-\$0) | \$2,653,252<br>(3.5%) |
| • Associated Research Funding | \$0<br>(\$0-\$66,026) | \$59,768,145<br>(80.0%) |
| **Undisclosed COIs (All categories)** | \$ 4,178<br>(\$227-\$62,564) | \$23,976,655<br>(24.3%) |
| • Total Direct Payments | \$ 1,153<br>(\$113-\$9,902) | \$3,964,536<br>(16.5%) |
| ○ General payments | \$992<br>(\$60-\$8,509) | \$3,769,344<br>(15.7%) |
| ▪ Food & beverage | \$191<br>(\$20-\$988) | \$195,191<br>(0.8%) |
| ▪ Others* | \$ 268<br>(\$0-\$6,810) | \$3,574,153<br>(14.9%) |
| ○ Direct research payment | \$0<br>(\$0-\$0) | \$197,942<br>(0.8%) |
| • Associated Research Funding | \$0<br>(\$0-\$35,416) | \$20,012,119<br>(83.5%) |

\* Other general payment includes consulting, honoraria, royalty or license, education, gifts, and travel and lodging.
Abbreviations: COI = conflict of interest; IQR = interquartile range

Among all medical professional societies, the guideline panel members of the American Academy of Dermatology had the highest general payments received (mean [IQR], $70,727 [$3,945-$544,211]), while panel members from the American Society of Anesthesiologists received the lowest general payments (mean [IQR], $62 [$58-$65]). More details about the identified COI by medical professional societies are reported in Supplemental Table 3.

While 15 (7.5%) authors with financial COIs on Open Payments disclosed all received payments, 108 (54.3%) did not disclose any payments (Supplemental Figure 1). Among the authors with undisclosed or underreported COIs (n=184), 58.7% of authors’ nondisclosures were for Direct Payments (4.9% general payments only, 53.8% combination of general payments and direct research payments), 5.4% for Associated Research Funding, and 35.9% for a combination of Direct payments and Associated Research funding (Figure 1).

**Figure 1.**
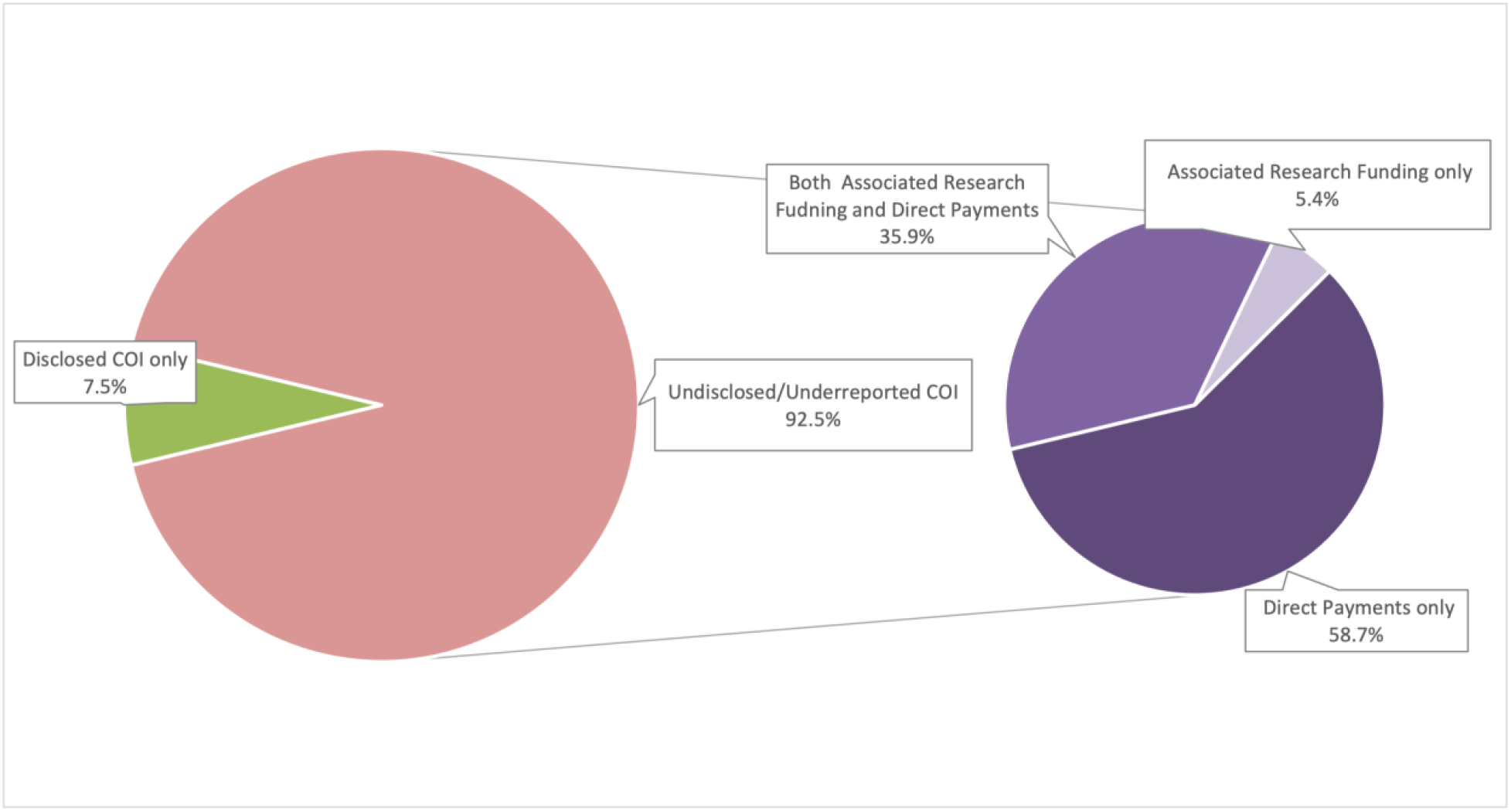
Types of Financial Conflict of Interest Under- and Undisclosed among U.S. Physician Authors of 2020 Clinical Practice Guidelines

## DISCUSSION

In our cross-sectional study of 2020 clinical practice guidelines that compared self-reported financial COIs with payments from industry reported to CMS through the Open Payments program, we found that financial COIs are common among U.S. physician guideline panel members and are often not disclosed accurately. Although the majority of guideline authors had financial relationships with industry, more than 90% did not completely disclose all financial COIs. These findings raise concerns about potential bias in the treatment recommendations developed by key medical professional societies in the United States.

The National Academy of Medicine recommends guideline panel chairs and co-chairs to not have any conflicts, and that only a minority of guideline authors should have a financial COI.^2^ However, consistent with prior research^11^, our analysis identified a majority of 2020 guidelines within our sample had panel chairs with COI, all of which inaccurately disclosed their COI. Moreover, for most guidelines, authors with financial COI comprised the majority of the panels. Our study demonstrates that even among more contemporary guideline panels, when professional organizations had the opportunity to scrutinize financial COIs among physicians who were being considered for panel membership, financial COIs were common and remain inaccurately disclosed. Because financial COIs create a risk that professional judgments or actions may be unduly influenced by secondary interests, our findings raise concerns about guidelines’ quality, reliability, and integrity.

Although a large proportion of the monetary value of financial COIs were associated with research activities through institutions, we found that authors were more likely to have undisclosed or underreported COIs for direct payments. Considering that direct payments could potentially have a greater impact on panel members’ decisions, more attention should be paid to such COIs. Certain medical professional societies also had higher rates of COIs, inaccurate disclosures, and greater values of payments received from the industry among their panel members, thus necessitating more rigorous action to be taken by those societies, perhaps with oversight from CMSS. Disclosure, assessment, and management of COIs is a process that requires consideration throughout the guideline development, particularly since relationships may change. Utilizing specific structured disclosure forms with closed-ended questions may improve the accuracy of COI disclosure.^9^ These forms should inquire about both active and inactive relationships with the industry ahead of the process of guideline development to ensure compliance with National Academy of Medicine recommendations. Additional detailed questions can further clarify the relevancy and extent of those financial relationships. Moreover, medical professional societies should evaluate the completeness of COI disclosure by comparing the self-reported COIs with data available on Open Payments. Thereafter, all COIs that potentially affect guideline development should be managed appropriately.

This study had certain limitations. First, although we included an eligible guideline from all the CMSS members, it was not feasible to include all the guidelines published by CMSS in 2020. Among those with multiple guidelines, we selected the ones with the largest number of authors to have an appropriate sample. Also, we included only physicians based in the U.S since other guideline authors would not have profiles on the Open Payments database. Second, data available on Open Payments, although frequently updated and verified by payment recipients, does not contain all the payments received and may not be fully accurate.^25^ Third, we attempted to characterize all payments from industry to physicians reported through the Open Payments program in the three years prior to guideline publication, in alignment with ICMJE disclosure requirements.^22^ However, our look back may be imprecise because exact dates for guidelines’ convening, which may have taken months to more than a year to finalize, and for guidelines’ first submission to a journal for consideration, were not consistently available. Lastly, we only considered the pharmaceutical and medical device industry-related financial COIs. Although other financial COIs and other types of COIs could influence the quality of clinical practice guidelines, Open Payments only records industry payments and does not contain data related to other COIs. Despite these limitations, our study included a wide range of contemporary clinical practice guidelines from different societies, making the findings more generalizable than those of similar studies.

## CONCLUSION

Financial COIs among U.S. physician authors of clinical practice guidelines are common and are often not disclosed accurately. Given the importance of clinical practice guidelines in both providing care to patients and guiding future research in medicine, these guidelines should be as accurate and unbiased as possible. The substantial COIs that exist among guideline authors and the inconsistencies between payments reported by industry and COI self-reported within the guidelines emphasized the need for implementing greater oversight and additional policies for disclosing and managing COIs in medical professional societies producing clinical practice guidelines to ensure their quality, reliability, and integrity.

## Data Availability

Relevant data are available on reasonable request from the corresponding author.

## Ethics statements

Ethical approval: Not required. Publicly available nonclinical datasets were used.

## Contributorship statement

MM, LG and JSR conceived of and designed the study. MM and LG collected the data. MM led the data analysis and drafted the first version of the manuscript. All authors reviewed and interpreted the data, read the manuscript and provided critical feedback for important intellectual content. JSR provided supervision. All authors approved the submission of the current version of the manuscript. The corresponding author attests that all listed authors meet authorship criteria and that no others meeting the criteria have been omitted.

## Competing interests

Drs. Mooghali, Ramachandran, and Ross currently receive research support through Yale University from Arnold Ventures. Dr. Ramachandran currently receives research support through Yale Law School from the Stavros Niarchos Foundation for a project focused on public R&D and manufacturing for enabling equitable access to medical technologies. She also serves as a consultant to the ReAct-Action on Antibiotic Resistance Strategic Policy Program based out of Johns Hopkins Bloomberg School of Public Health, which is funded by the Swedish International Development and Cooperation Agency (Sida). Dr. Ross currently receives research support through Yale University from Johnson and Johnson to develop methods of clinical trial data sharing, from the Medical Device Innovation Consortium as part of the National Evaluation System for Health Technology (NEST), from the Food and Drug Administration for the Yale-Mayo Clinic Center for Excellence in Regulatory Science and Innovation (CERSI) program (U01FD005938), from the Agency for Healthcare Research and Quality (R01HS022882), and from the National Heart, Lung and Blood Institute of the National Institutes of Health (NIH) (R01HS025164, R01HL144644); in addition, Dr. Ross is an expert witness at the request of Relator’s attorneys, the Greene Law Firm, in a qui tam suit alleging violations of the False Claims Act and Anti-Kickback Statute against Biogen Inc.

## Funding

No external grants or funds were used to support this project.

## Data Sharing statement

Relevant data are available on reasonable request from the corresponding author.

