## Supplemental files for "Financial Conflicts of Interest among U.S. Physician Authors of 2020 Clinical Practice Guidelines: A Cross-Sectional Study"

**List of Supplemental Tables and Figures**

Supplemental Table 1 - 2020 Clinical Practice Guidelines published by the Council of Medical Specialty Societies

Supplemental Table 2 - Characteristics of Guideline Authors of 2020 Clinical Practice Guidelines published by the Council of Medical Specialty Societies

Supplemental Table 3 - Financial Conflict of Interests among U.S. Physician Authors of 2020 Clinical Practice Guidelines, Stratified by Medical Professional Societies

Supplemental Figure 1 – Conflict of Interest Among U.S. Physician Authors of 2020 Clinical Practice Guidelines, Stratified by Proportions of Undisclosed/Total Conflict of Interest

*Supplemental Table 4 - 2020 Clinical Practice Guidelines published by the Council of Medical Specialty Societies*

| Medical Professional Society | Guideline | Total number of listed authors | Number of U.S.-based physicians listed authors |
|---|---|---|---|
| American Academy of Allergy, Asthma & Immunology | Anaphylaxis—a 2020 practice parameter update, systematic review, and Grading of Recommendations, Assessment, Development and Evaluation (GRADE) analysis[1] | 17 | 15 |
| American Academy of Dermatology | Joint American Academy of Dermatology - National Psoriasis Foundation guidelines of care for the management of psoriasis with systemic nonbiologic therapies[2] | 34 | 31 |
| American Academy of Neurology | Practice Guideline: Treatment for Insomnia and Disordered Sleep Behavior in Children and Adolescents with Autism Spectrum Disorder[3] | 26 | 17 |
| American College of Cardiology | 2020 AHA/ACC Guideline for the Diagnosis and Treatment of Patients with Hypertrophic Cardiomyopathy[4] | 19 | 15 |
| American College of Emergency Physicians | Clinical Policy: Critical Issues Related to Opioids in Adult Patients Presenting to the Emergency Department[5] | 7 | 7 |
| American College of Physicians | Testosterone Treatment in Adult Men With Age-Related Low Testosterone: A Clinical Guideline From the American College of Physicians[6] | 5 | 4 |
| American College of Rheumatology | 2020 American College of Rheumatology Guidelines for the Management of Reproductive Health in Rheumatic and Musculoskeletal Diseases[7] | 36 | 24 |
| American Gastroenterological Association | AGA Clinical Practice Guidelines on the Gastrointestinal Evaluation of Iron Deficiency Anemia[8] | 7 | 7 |
| American Society of Anesthesiologists | Practice Guidelines for Central Venous Access 2020: An Updated Report by the American Society of Anesthesiologists Task Force on Central Venous Access[9] | 7 | 6 |
| American Society of Clinical Oncology | Metastatic Pancreatic Cancer: ASCO Guideline Update[10] | 19 | 16 |
| American Society of Colon and Rectal Surgeons | The American Society of Colon and Rectal Surgeons Clinical Practice Guidelines for the Surgical Management of Crohn's Disease[11] | 10 | 10 |
| American Society of Hematology | American Society of Hematology 2020 guidelines for treating newly diagnosed acute myeloid leukemia in older adults[12] | 23 | 14 |

| | | | |
|---|---|---|---|
| American Society for Radiation Oncology | Radiation Therapy for Small Cell Lung Cancer: An ASTRO Clinical Practice Guideline[13] | 17 | 14 |
| American Society for Reproductive Medicine | Evidence-based treatments for couples with unexplained infertility: a guideline[14] | 15 | 13 |
| American Thoracic Society | Initiating Pharmacologic Treatment in Tobacco-Dependent Adults: An Official American Thoracic Society Clinical Practice Guideline[15] | 30 | 10 |
| American Urological Association | Microhematuria: AUA/SUFU Guideline[16] | 15 | 14 |
| Infectious Diseases Society of America | Clinical Practice Guidelines by the IDSA: 2020 Guideline on the Diagnosis and Management of Babesiosis[17] | 14 | 10 |
| American Academy of Family Physicians | Nonpharmacologic and Pharmacologic Management of Acute Pain From Non–Low Back, Musculoskeletal Injuries in Adults: A Clinical Guideline From the American College of Physicians and American Academy of Family Physicians[18] | 6 | 6 |
| Society of Critical Care Medicine | Surviving Sepsis Campaign International Guidelines for Management of Septic Shock and Sepsis-Associated Organ Dysfunction in Children[19] | 51 | 25 |
| Society for Vascular Surgery | Society for Vascular Surgery (SVS) and Society of Thoracic Surgeons (STS) reporting standards for type B aortic dissections[20] | 13 | 12 |

*Supplemental Table 5 - Characteristics of Guideline Authors of 2020 Clinical Practice Guidelines published by the Council of Medical Specialty Societies*

| Characteristics | N (%)<br>(n=371) |
|---|---|
| Gender | |
| • Male | 221 (59.6%) |
| • Female | 145 (39.1%) |
| • Unclear | 5 (1.3%) |
| Rank | |
| • Professor | 174 (46.9%) |
| • Associate Professor | 87 (23.5%) |
| • Assistant Professor | 44 (11.9%) |
| • Other / Not Reported | 66 (17.8%) |
| Location | |
| • United States | 309 (83.3%) |
| • Canada (No profile on Open Payments) | 28 (7.5%) |
| • Other Countries (No profile on Open Payments) | 34 (9.2%) |
| Degree | |
| • MD/DO/MBBS | 318 (85.7%) |
| • Non-MD/DO/MBBS | 53 (14.3%) |
| Profile on Open Payments | |
| • No available profile - Excluded from the analysis | 101 (27.2%) |
| • Available Profile - Included in the analysis | 270 (72.8%) |
| Conflict of Interest declared in the guideline | |
| • Authors who declared industry-related COIs in the guidelines | 129 (34.8%) |
| • Authors who did NOT declare any industry-related COIs in the guidelines | 242 (65.2%) |

Abbreviations: COI = conflict of interest; DO = Doctor of Osteopathic Medicine; MBBS = Bachelor of Medicine, Bachelor of Surgery; MD = Doctor of Medicine.

*Supplemental Table 6 - Financial Conflict of Interests among U.S. Physician Authors of 2020 Clinical Practice Guidelines, Stratified by Medical Professional Societies*

| Medical Professional Society | Number of Included Authors | Number of authors with COI (%) | General payments received, Mean (IQR) | Direct research payments received, Mean (IQR) | Associated research funding received, Mean (IQR) |
|---|---|---|---|---|---|
| American Academy of Allergy, Asthma & Immunology | 15 | 13 (86.7%) | $32,119 ($4,933-$68,247) | $0 ($0-$2,403) | $2,500 (0-$72,166) |
| American Academy of Dermatology | 31 | 26 (83.9%) | $70,727 ($3,945-$544,211) | $19,333 ($0-$47,124) | $140,916 ($0-$1,735,916) |
| American Academy of Neurology | 17 | 12 (70.6%) | $1,128 ($176-$6,002) | $0 ($0-$0) | $0 ($0-$11,836) |
| American College of Cardiology | 15 | 11 (73.3%) | $439 ($60-$11,982) | $0 ($0-$0) | $0 ($0-$114,716) |
| American College of Emergency Physicians | 7 | 2 (28.6%) | $533 ($280-$787) | $0 ($0-$0) | $0 ($0-$0) |
| American College of Physicians | 4 | 1 (25.0%) | $239 ($239-$239) | $0 ($0-$0) | $0 ($0-$0) |
| American College of Rheumatology | 24 | 16 (66.7%) | $3,180 ($91-$34,226) | $0 ($0-$1,097) | $25,423 ($0-$221,056) |
| American Gastroenterological Association | 7 | 3 (42.9%) | $188 ($118-$315) | $0 ($0-$0) | $0 ($0-$0) |
| American Society of Anesthesiologists | 6 | 2 (33.3%) | $62 ($58-$65) | $0 ($0-$0) | $0 ($0-$0) |
| American Society of Clinical Oncology | 16 | 14 (87.5%) | $20,332 ($2,126-$49,587) | $1,315 ($0-$4,213) | $588,530 ($203,102-$2,730,253) |
| American Society of Colon and Rectal Surgeons | 10 | 10 (100.0%) | $18,990 ($12,137-$69,903) | $0 ($0-$0) | $266 ($0-$30,962) |
| American Society of Hematology | 14 | 13 (92.9%) | $11,239 ($1,286-$133,932) | $1,221 ($0-$27,779) | $477,734 ($222,642-$803,713) |
| American Society for Radiation Oncology | 14 | 12 (85.7%) | $6,190 ($2,248-$35,673) | $0 ($0-$0) | $0 ($0-$74,438) |
| American Society for Reproductive Medicine | 13 | 11 (84.6%) | $834 ($54-$5,444) | $0 ($0-$0) | $0 ($0-$0) |
| American Thoracic Society | 10 | 6 (60.0%) | $445 ($249-$11,657) | $0 ($0-$0) | $0 ($0-$0) |
| American Urological Association | 14 | 13 (92.9%) | $8,853 ($1,120-$28,184) | $0 ($0-$0) | $0 ($0-$10,000) |

| | | | | | |
|---|---|---|---|---|---|
| Infectious Diseases Society of America | 10 | 5 (50.0%) | $133 ($70-$12,757) | $0 ($0-$0) | $0 ($0-$36,825) |
| American Academy of Family Physicians | 6 | 2 (33.3%) | $2,482 ($1,372-$3,539) | $0 ($0-$0) | $0 ($0-$0) |
| Society of Critical Care Medicine | 25 | 15 (60.0%) | $291 ($6 - $3,637) | $0 ($0-$0) | $0 ($0-$6,917) |
| Society for Vascular Surgery | 12 | 12 (100.0%) | $28,714 ($18,066-$85,445) | $0 ($0-$0) | $54,800 ($6,155-$350,983) |

Abbreviations: COI = conflict of interest; IQR = interquartile range.

*Supplemental Figure 2 – Conflict of Interest Among U.S. Physician Authors of 2020 Clinical Practice Guidelines, Stratified by Proportions of Undisclosed/Total Conflict of Interest*

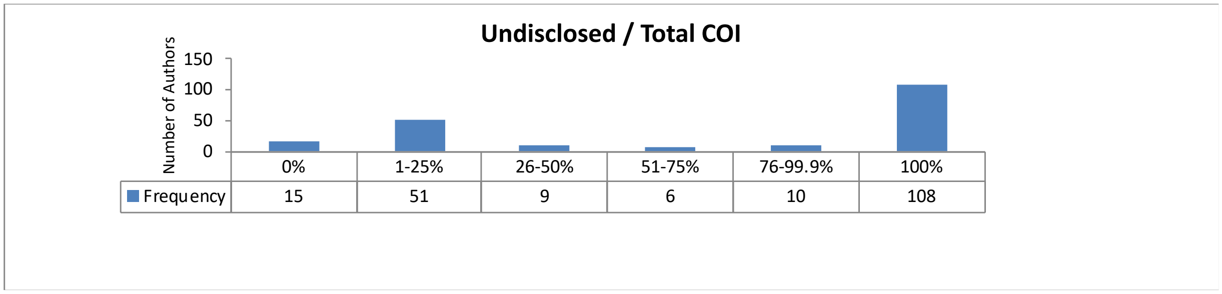


Abbreviations: COI = conflict of interest.
